# Snapshot: Pregnancy-Related Stroke

**DOI:** 10.1101/2025.05.12.25327067

**Authors:** Ming Zheng

## Abstract

Pregnancy-related stroke is a previously overlooked risk factor. Addressing this issue is of crucial importance, not just because pregnancy-related disorder affects women of reproductive age, but also because it is often preventable. However, it is difficult to determine whether these disorders in the brief nine months of pregnancy may lead to later stroke risk, because of the difficulties in the systematic collection of long-time follow-up data and the control for confounding factors and potential bias. To address this issue, this study sought to conduct a Mendelian randomization (MR) study and discovered preeclampsia, HELLP syndrome, gestational hypertension and diabetes as major pregnancy-related disorders that pose increased risks of stroke. This study is meant to provide a comprehensive landscape of stroke risks across different pregnancy complications, most of which are preventable and treatable, thus generating many testable candidates of strong associations. From this perspective, these associations may serve as a pragmatic road map for researchers who aim to investigate pregnancy-related stroke, for clinicians who wish to improve maternal care to prevent stroke, and for patients who need to take appropriate precautionary measures or behavior modifications to reduce stroke risk.

## Introduction

Globally, around 85% of women experience pregnancy at some point in their lives, with approximately 140 million women becoming pregnant each year.^1^ During the roughly 40-week gestation period, the complex process of pregnancy is mediated by the placenta that allows the fetus to ‘plug into’ the mother’s cardiovascular system. Pregnancy imposes significant physiological requirements on the mother as she adapts to accommodate the development of fetus. These physiological accommodations, including increases in blood volume, metabolic rate, oxygen consumption, and adjustments in immune function,^2^ make women more vulnerable to various health complications both during pregnancy and in the postpartum period.

It has been reported that pregnancy-associated stroke is the most common cause of mortality and disability following pregnancy.^3^ Stroke in pregnancy is linked to several pregnancy-related disorders, including preeclampsia and gestational diabetes, which elevate the risk of both arterial and venous thrombosis.^4^ However, despite the growing recognition of these associations, pregnancy-related disorders have been underexplored in stroke research.

Until recently, there has been sporadic reporting of increasing stroke incidence in young women of reproductive age,^5,6^ and women with pregnancy-related disorders may have an increased risk of stroke for many years after childbearing.^7^ This underscores the urgent need for further research to understand the causal role of pregnancy-related disorders on stroke risk. However, it is still difficult to determine whether the physiological changes during the relatively short duration of pregnancy contribute to an increased risk of stroke later in life. Long-term follow-up data, which are essential for understanding this relationship, are difficult to collect systematically. Additionally, controlling for confounding factors, such as preexisting health conditions and lifestyle factors, introduces further complexity.

## Methods and Materials

### Study design and data sources

This study performed a two-sample Mendelian randomization (MR) analysis to assess the potential causal effects of genetically determined pregnancy-related disorders on the risk of stroke. Summary-level association statistics for stroke (all-cause and subtype-specific) were obtained from the largest available genome-wide association study (GWAS) meta-analysis of European-ancestry participants (N = 521,612), which encompasses ischemic stroke (IS), hemorrhagic stroke, and IS subtypes (cardioembolic, large-artery, small-vessel).^8^ Summary statistics for major pregnancy-related disorders—such as preeclampsia, HELLP syndrome, gestational hypertension, and gestational diabetes—were drawn from independent GWAS meta-analyses.^9^

### Selection of genetic instruments and the exposure and outcome data

Independent single-nucleotide polymorphisms (SNPs) strongly associated with each pregnancy-related disorder were selected as instrumental variables (IVs) using a *p*-value <5×10^−7^ and an independent inheritance with a minimal level of linkage disequilibrium (LD) of r^2^ <0.001. Instrumental strength was quantified using the *F*-statistic, with an *F*-statistic >10 being considered sufficiently informative.^10^ Finally, 57 SNPs were selected with an average *F*-statistic of 41.4 (range 25.3-234.7), thus representing sufficient instrumental strength; variances explained ranged from 1.5% to 24.4% (average =6.6%) for different pregnancy-related disorders.

### Mendelian randomization analysis

Causal effect estimates with sensitivity analyses were primarily obtained using four MR methods, including simple mode, weighted median, weighted mode, and inverse-variance weighting (IVW). The causal estimate was calculated by log-odds ratio. MR pleiotropy residual sum and outlier (MR-PRESSO) test was conducted to correct the horizontal pleiotropy when the MR-PRESSO global test *p*-value was <0.05.^11^

### Software for statistical analysis

All analyses were conducted in R 3.6.1 using the packages TwoSampleMR (v0.5.6) for core MR methods, MRPRESSO (v1.0) for pleiotropy correction, and ld_clump in ieugwasr (v0.1.5) for LD clumping on SNP instruments. LD reference used the 1000 Genomes Phase 3 European panel.

## Results

MR analysis was performed to estimate the risk of stroke (outcome) following pregnancy-related disorders (exposure) (**Figure. 1A**). As shown in **Figure. 1B**, increased risk of stroke was significantly associated with different pregnancy-related disorders, including gestational hypertension, preeclampsia, HELLP syndrome (**H**emolysis, **E**levated **L**iver enzymes, **L**ow **P**latelets), and gestational diabetes (adjusted *P*_IVW_ <0.05). Intriguingly, this study also found excessive vomiting in pregnancy to be associated with increased stroke risk (adjusted *P*_IVW_ <0.05; **Figure. 1B**), indicating excessive vomiting in pregnancy as a possible sign of increased stroke risk.

**Figure 1.**
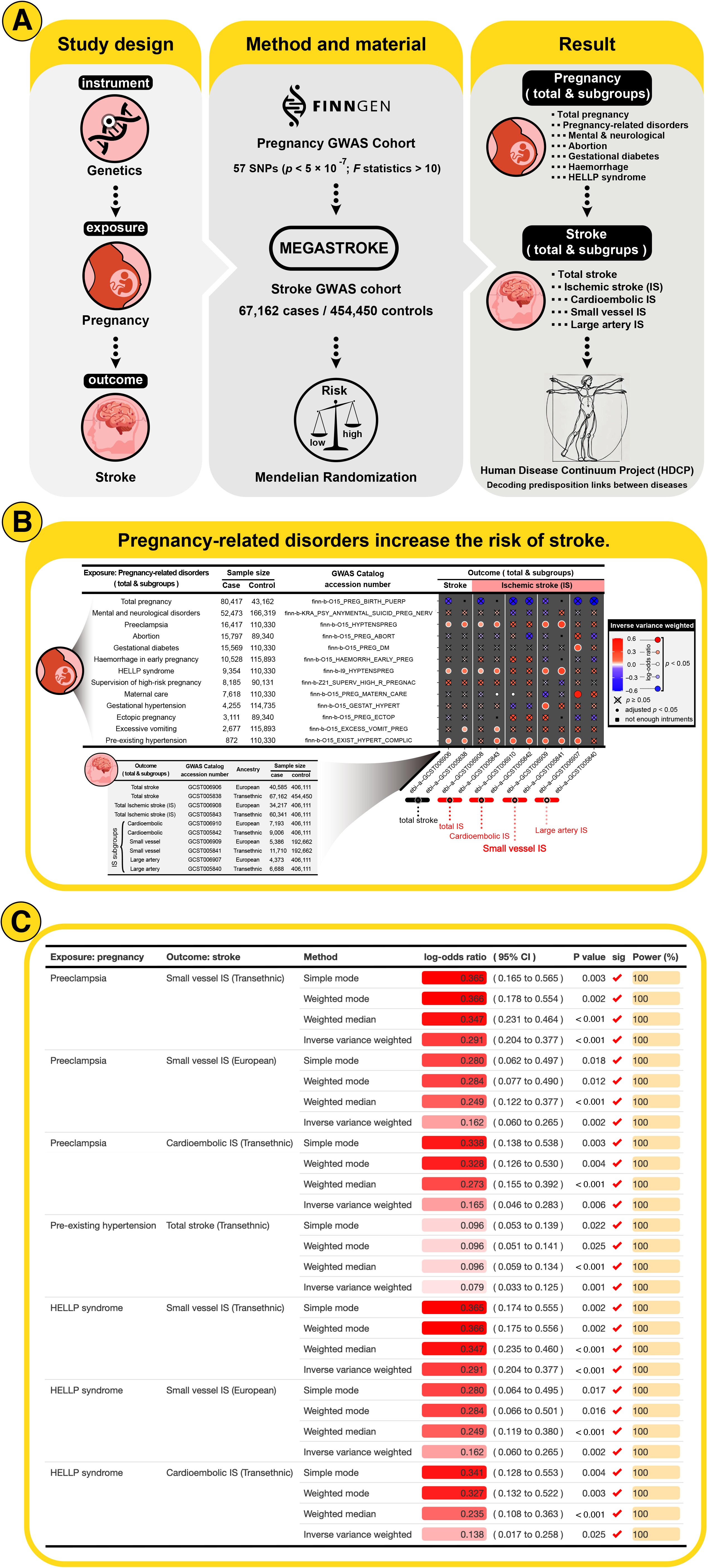
Mendelian randomization study of pregnancy-related disorders and stroke. **(A)** The graphical illustration of the Mendelian randomization (MR) study. The MR analysis was conducted using the genetic instrument to estimate the causal effect of pregnancy-related disorders exposure on stroke risk. This MR analysis tested the 57 genetic variants of pregnancy-related disorders against stroke in genome-wide association study (GWAS) cohorts to evaluate the stroke risk following pregnancy-related disorders. **(B)** MR results show the causal effect of pregnancy-related disorders on stroke susceptibility. MR estimated causal effects were calculated using the inverse variance weighted test, with horizontal pleiotropy corrected when MR pleiotropy residual sum and outlier (MR-PRESSO) Global test *p* <0.05. The causal estimates were shown by heatmap, with the dot color and size representing the log-odds ratios. A positive log-odds ratio indicates that pregnancy-related disorders are causally associated with an increased risk of stroke. The “×” symbol represents adjusted *p* ≥ 0.05; and, the dot with a white outer ring symbol represents false discovery rate (FDR)-adjusted *p* <0.05. **(C)** Causal estimates of genetically determined pregnancy-related disorders on stroke risk. The log-odds ratios were analyzed by three different MR tests: simple mode, weighted mode, weighted median, and inverse variance weighted (IVW). The significance of *p* <0.05 was indicated by “✓” symbol. A power calculation was performed to test whether the MR analysis was adequately powered to detect the causal effect. Power (%) = the statistical power of MR analysis to detect the corresponding causal effect at α =0.05, represented by bar length. The MR causal effect was shown by the color gradient.

Next, the above MR results were validated using other MR methods: simple mode, weighted median, and weighted mode; and the statistical power was calculated by the power calculation. The most significant causality of ischemic stroke (IS) was found in preeclampsia and HELLP syndrome, and the results were consistent across different MR tests (*P*_simple mode_ <0.05, *P*_weighted mode_ <0.05, *P*_weighted median_ <0.05, *P*_IVW_ <0.05; power =100%; **Figure. 1C**). Of note, this study found preeclampsia and HELLP syndrome to be associated with specific stroke subtypes of cardioembolic and small-vessel IS. In general, there was a positive trend toward increased stroke risk following different pregnancy-related disorders, including preeclampsia, HELLP syndrome, gestational hypertension and diabetes (**Figure. 1C; Supplemental Figure. 1**).

## Discussion

Human pregnancy is relatively long in supporting fetal brain and body development. This extended gestation period places a significant strain on the mother’s body, requiring considerable cardiovascular anatomical adjustments. For example, cardiac output—the amount of blood pumped by the heart per minute—increases by nearly 50%. This surge in output ensures that both maternal tissues and fetus receive sufficient blood flow. To maintain this, the heart enlarges slightly, and blood volume increases dramatically.^12^ These maternal cardiovascular alterations can put a strain on the mother’s heart and circulatory system, raising the risk of complications like hypertension or stroke. Human pregnancy reflects a delicate evolutionary trade-off placed on the maternal body, where essential adaptations to support fetal development also pose cardiovascular risks to the mother.

Pregnancy-related stroke represents a critical and under-examined area in the intersection of obstetric care and neurological outcomes. While pregnancy is a unique and inherently risky physiological state, stroke, a disease typically associated with older age, has been increasingly recognized as a significant cause of morbidity and mortality among young women, particularly those with pregnancy-related disorders. Stroke occurring in the postpartum period always results in rehospitalization, with a case fatality rate of 7.4%.^4^ This study makes a crucial contribution by linking pregnancy complications—such as preeclampsia, HELLP syndrome, gestational hypertension, and gestational diabetes—to an increased long-term risk of stroke, particularly ischemic stroke. By employing Mendelian randomization (MR) and genomic data, the study strengthens causal inferences that these pregnancy-related disorders have lasting vascular implications, potentially predisposing women to cardiovascular events long after childbirth.

This study underscore a pressing public health concern: the long-term vascular risks associated with pregnancy-related disorders. The association between preeclampsia, HELLP syndrome, gestational hypertension, and gestational diabetes with an elevated stroke risk is compelling. These disorders are well-established as significant contributors to maternal morbidity during pregnancy, and they have now been implicated in stroke pathogenesis, not just during pregnancy but also in the postpartum period and beyond.

Preeclampsia, characterized by hypertension and proteinuria, is associated with endothelial dysfunction, thrombophilia, and a heightened risk of arterial damage. HELLP syndrome, a severe form of preeclampsia, involves hemolysis, elevated liver enzymes, and low platelet count, often precipitating a more aggressive vascular and thrombotic response. Both conditions are known to increase the risk of ischemic stroke during pregnancy and the early postpartum period. Importantly, the study highlights how these pregnancy-related disorders may predispose women to specific subtypes of ischemic stroke, such as cardioembolic and small-vessel strokes. This distinction is critical as it suggests that pregnancy-related strokes may have a distinct pathophysiology, requiring tailored prevention and management strategies.

Gestational hypertension and diabetes, both of which are increasingly common due to rising rates of obesity and metabolic disorders, further compound stroke risk. These conditions lead to both endothelial injury and pro-inflammatory states that contribute to vascular damage and thrombosis. Furthermore, the identification of excessive vomiting in pregnancy (hyperemesis gravidarum) as a potential stroke risk factor is an intriguing finding that warrants further investigation. While the precise mechanism remains unclear, it suggests that extreme physiological stress during pregnancy may act as a surrogate marker for more systemic vascular complications.

### Strengths and Limitations of the Study

One of the key strengths of this study is its use of Mendelian randomization (MR), which leverages genetic variation as a natural experiment to infer causal relationships. This methodological approach addresses the challenge of confounding factors that often undermine observational studies, such as pre-existing cardiovascular risk factors or other lifestyle factors (e.g., smoking or diet). The large sample size (521,612 participants) and the genetic instruments employed (57 SNPs) enhance the power and reliability of the findings. The MR-PRESSO test further mitigates the risk of pleiotropy—where genetic instruments affect multiple traits—by adjusting for outliers and ensuring that the associations are not driven by indirect genetic effects.

However, the study is not without limitations. The relatively small sample size of specific stroke subtypes, such as large-artery stroke, may limit the ability to detect significant associations in these groups. Larger studies with more granular subtype analysis are needed to confirm the generalizability of the findings across all stroke phenotypes.

### Clinical Relevance of the Study

This study is meant to provide a comprehensive landscape of stroke risks across different pregnancy complications, these associations may serve as a pragmatic road map for researchers who aim to investigate pregnancy-related stroke, for clinicians who wish to improve maternal care to prevent stroke, and for patients who need to take appropriate precautionary measures or behavior modifications to reduce stroke risk. Thus, understanding the long-term cardiovascular risks associated with pregnancy-related disorders should become an integral part of maternal health care. Preeclampsia, HELLP syndrome, gestational hypertension, and diabetes should be considered not only as pregnancy-related concerns but also as risk factors for future cardiovascular disease, including stroke.

A comprehensive approach to stroke prevention in women with a history of pregnancy-related disorders should be instituted, focusing on both preconception counseling and postpartum care. Women with a history of these conditions should receive tailored follow-up care that includes regular cardiovascular monitoring, including blood pressure checks, lipid profile assessments, and screening for diabetes. Additionally, lifestyle modifications—such as weight management, exercise, smoking cessation, and dietary changes—should be emphasized as part of stroke prevention strategies. Clinicians should be particularly vigilant in the postpartum period, as the risk of stroke is highest in the immediate weeks following childbirth. Timely recognition of symptoms, early intervention, and the avoidance of premature discharge are key in preventing devastating outcomes.

Furthermore, patients themselves should be educated about their elevated stroke risk and encouraged to take proactive steps in monitoring their health. This includes understanding the importance of regular blood pressure checks, avoiding smoking and excessive alcohol consumption, and being aware of the signs and symptoms of stroke. Educating patients to seek immediate medical attention in the event of a stroke-like symptom could significantly reduce morbidity and mortality associated with pregnancy-related strokes.

On a broader scale, this study calls attention to the growing need for targeted public health interventions aimed at improving maternal health. Pregnancy-related disorders are disproportionately experienced by women in low-resource settings, where access to high-quality maternal care is limited. This study supports the idea that improving prenatal care, monitoring at-risk pregnancies, and ensuring adequate postnatal follow-up could not only reduce maternal mortality but also prevent long-term cardiovascular complications, including stroke. Efforts to reduce health disparities in maternal care are essential in mitigating the public health burden of pregnancy-related strokes.

Additionally, there is a need for more research into the pathophysiology of pregnancy-related stroke. Understanding the genetic and environmental factors that contribute to these conditions could lead to more effective prevention strategies. Future studies should focus on larger, multi-center cohorts, with a more detailed analysis of stroke subtypes, to further refine the associations between pregnancy complications and stroke risk.

In future studies, integrating imaging phenotypes—such as magnetic resonance imaging (MRI) measures of cerebral vessel integrity—into longitudinal pregnancy cohorts of women before, during, and after pregnancy could chart the evolution of imaging signatures that bridge acute pregnancy disorders and chronic cerebrovascular disease. This would allow for further validation of the genomic prediction of pregnancy-related stroke risks. Moreover, extending analyses to diverse ancestry groups is essential to ensure global applicability and equity in stroke prevention.

### Implications for Human Disease Continuum Project (HDCP)

Integrating these findings into the framework of the Human Disease Continuum Project (HDCP)^13^ underscores how acute, pregnancy-related vascular insults can seed a trajectory of lifelong cerebrovascular risk. The HDC project conceptualizes human health as a dynamic spectrum, in which transient physiological perturbations—such as those imposed by pregnancy—may initiate pathophysiological cascades that manifest decades later. By applying Mendelian randomization to quantify the causal impact of preeclampsia, HELLP syndrome, gestational hypertension, gestational diabetes, and hyperemesis gravidarum on subsequent stroke risk, this study fills a critical gap in this continuum: the transition from reproductive-age vascular stressors to chronic cerebrovascular disease.

Firstly, the demonstration that genetically instrumented pregnancy disorders confer elevated long-term stroke risk provides empirical support for the principle of human disease continuum (HDC) that early-life exposures can have durable, latent effects. In the HDC model, each ‘hit’ against vascular integrity—whether environmental, genetic, or physiological—adds to cumulative damage. Pregnancy represents a potent, physiological “hit,” heightening hemodynamic load and endothelial stress. The robust associations, particularly for preeclampsia and HELLP syndrome with cardioembolic and small-vessel stroke subtypes, map precisely onto HDC predictions: severe vascular insults accelerate progression toward overt cerebrovascular pathology.

Secondly, situating pregnancy-related stroke within the HDC framework highlights opportunities for early intervention. Whereas stroke prevention efforts traditionally focus on mid-life or later risk factor control, the results advocate for a life-course approach beginning at—or even before—pregnancy. Identification of women with high genetic liability to pregnancy complications could inform preconception counseling, intensive monitoring during gestation, and structured postpartum cardiovascular follow-up. Embedding these strategies within the HDC project’s preventative architecture would enable targeted “course corrections” before irreversible vascular damage accrues.

Thirdly, the HDC paradigm emphasizes the shared mechanistic underpinnings linking disparate conditions across the lifespan. The finding that hyperemesis gravidarum—a condition not classically associated with vascular pathology—also predicts stroke risk suggests unrecognized vascular vulnerabilities may be unmasked by systemic stress. Investigating the overlap in genetic and molecular pathways among preeclampsia, hyperemesis, and later cerebrovascular injury could reveal novel intervention points, aligning with the HDC aim of uncovering common nodes in the disease network.

By demonstrating causal links between pregnancy-related disorders and long-term stroke risk, this study not only advances obstetric neurology but also enriches the Human Disease Continuum project’s life-course perspective. Recognizing pregnancy as a pivotal window into future cerebrovascular health invites a reorientation of research and clinical practice toward earlier, more personalized, and mechanism-based interventions—hallmarks of the HDC vision for a truly integrated model of human disease prevention.

## Conclusion

This study adds significantly to the understanding of pregnancy-related stroke by demonstrating that disorders like preeclampsia, HELLP syndrome, gestational hypertension, and diabetes are associated with increased stroke risk, both during and after pregnancy. Given the preventability of many pregnancy-related complications, improving maternal health care and ensuring long-term follow-up for women with these disorders is critical in reducing the long-term stroke burden. This research also highlights the need for continued investigation into the underlying mechanisms that link pregnancy-related disorders with cerebrovascular events, with the ultimate goal of improving stroke prevention and management in women.

## Data Availability

All data produced in the present study are available upon reasonable request to the author.

## Nonstandard Abbreviations and Acronyms

IS: ischemic stroke
MR: Mendelian randomization
MR-PRESSO: Mendelian randomization pleiotropy residual sum and outlier
GWAS: genome-wide association study
SNP: single nucleotide polymorphism
LD: linkage disequilibrium

## Declarations

## Ethical Approval and Consent to participate

No human subjects were directly involved in this study. All the data used in this study was derived from existing de-identified biological samples from prior studies. Thus, ethical and patient consent was not required in this study.

## Acknowledgements

I would like to acknowledge the participants and investigators of the FinnGen project and the MEGASTROKE (International Stroke Genetics Consortium).

## Availability of supporting data

The data that support the findings of this study will be available from the corresponding author upon reasonable request.

## Authors’ contributions

M.Z. designed the study, developed the method, conducted data analysis, and wrote the manuscript. M.Z. supervised this project and is responsible for the overall content.

