## Supplemental Figure. 1 for "Snapshot: Pregnancy-Related Stroke"

A

| Exposure: pregnancy | Outcome: stroke | Method | log-odds ratio | ( 95% CI ) | P value | sig | Power (%) |
|---|---|---|---|---|---|---|---|
| Preeclampsia | Total stroke (Transethnic) | Simple mode | 0.104 | ( -0.015 to 0.224 ) | 0.102 | ✖ | 100 |
| | | Weighted mode | 0.118 | ( 0.011 to 0.224 ) | 0.042 | ✔ | 100 |
| | | Weighted median | 0.107 | ( 0.053 to 0.162 ) | < 0.001 | ✔ | 100 |
| | | Inverse variance weighted | 0.105 | ( 0.054 to 0.156 ) | < 0.001 | ✔ | 100 |
| Preeclampsia | Total stroke (European) | Simple mode | 0.079 | ( -0.030 to 0.188 ) | 0.167 | ✖ | 100 |
| | | Weighted mode | 0.088 | ( -0.021 to 0.196 ) | 0.125 | ✖ | 100 |
| | | Weighted median | 0.077 | ( 0.024 to 0.130 ) | 0.005 | ✔ | 100 |
| | | Inverse variance weighted | 0.064 | ( 0.027 to 0.100 ) | 0.001 | ✔ | 100 |
| Preeclampsia | Ischemic stroke (Transethnic) | Simple mode | 0.061 | ( -0.074 to 0.197 ) | 0.385 | ✖ | 100 |
| | | Weighted mode | 0.083 | ( -0.047 to 0.212 ) | 0.224 | ✖ | 100 |
| | | Weighted median | 0.068 | ( 0.006 to 0.131 ) | 0.031 | ✔ | 100 |
| | | Inverse variance weighted | 0.107 | ( 0.054 to 0.160 ) | < 0.001 | ✔ | 100 |
| Preeclampsia | Cardioembolic IS (European) | Simple mode | 0.242 | ( -0.030 to 0.513 ) | 0.093 | ✖ | 100 |
| | | Weighted mode | 0.246 | ( 0.019 to 0.473 ) | 0.044 | ✔ | 100 |
| | | Weighted median | 0.172 | ( 0.045 to 0.299 ) | 0.008 | ✔ | 100 |
| | | Inverse variance weighted | 0.137 | ( 0.024 to 0.249 ) | 0.018 | ✔ | 100 |
| Pre-existing hypertension | Total stroke (European) | Simple mode | 0.073 | ( 0.020 to 0.126 ) | 0.073 | ✖ | 92 |
| | | Weighted mode | 0.073 | ( 0.022 to 0.125 ) | 0.069 | ✖ | 92 |
| | | Weighted median | 0.072 | ( 0.032 to 0.113 ) | 0.001 | ✔ | 91 |
| | | Inverse variance weighted | 0.063 | ( 0.029 to 0.096 ) | < 0.001 | ✔ | 81 |
| Pre-existing hypertension | Small vessel IS (Transethnic) | Simple mode | 0.159 | ( 0.055 to 0.262 ) | 0.058 | ✖ | 99 |
| | | Weighted mode | 0.158 | ( 0.065 to 0.252 ) | 0.045 | ✔ | 99 |
| | | Weighted median | 0.158 | ( 0.085 to 0.231 ) | < 0.001 | ✔ | 98 |
| | | Inverse variance weighted | 0.134 | ( 0.072 to 0.196 ) | < 0.001 | ✔ | 93 |
| Pre-existing hypertension | Small vessel IS (European) | Simple mode | 0.135 | ( 0.013 to 0.256 ) | 0.119 | ✖ | 67 |
| | | Weighted mode | 0.134 | ( 0.012 to 0.257 ) | 0.120 | ✖ | 67 |
| | | Weighted median | 0.130 | ( 0.033 to 0.228 ) | 0.009 | ✔ | 64 |
| | | Inverse variance weighted | 0.124 | ( 0.040 to 0.209 ) | 0.004 | ✔ | 60 |
| Pre-existing hypertension | Large artery IS (Transethnic) | Simple mode | 0.133 | ( 0.013 to 0.252 ) | 0.118 | ✖ | 76 |
| | | Weighted mode | 0.133 | ( 0.019 to 0.246 ) | 0.107 | ✖ | 76 |
| | | Weighted median | 0.128 | ( 0.035 to 0.220 ) | 0.007 | ✔ | 72 |
| | | Inverse variance weighted | 0.103 | ( 0.026 to 0.179 ) | 0.008 | ✔ | 53 |
| Pre-existing hypertension | Large artery IS (European) | Simple mode | 0.208 | ( 0.054 to 0.362 ) | 0.077 | ✖ | 94 |
| | | Weighted mode | 0.207 | ( 0.067 to 0.347 ) | 0.062 | ✖ | 94 |
| | | Weighted median | 0.193 | ( 0.080 to 0.306 ) | 0.001 | ✔ | 90 |
| | | Inverse variance weighted | 0.164 | ( 0.071 to 0.256 ) | 0.001 | ✔ | 77 |
| Pre-existing hypertension | Ischemic stroke (Transethnic) | Simple mode | 0.108 | ( 0.057 to 0.159 ) | 0.054 | ✖ | 100 |
| | | Weighted mode | 0.108 | ( 0.052 to 0.163 ) | 0.063 | ✖ | 100 |
| | | Weighted median | 0.107 | ( 0.061 to 0.153 ) | < 0.001 | ✔ | 100 |
| | | Inverse variance weighted | 0.105 | ( 0.071 to 0.140 ) | < 0.001 | ✔ | 100 |
| Pre-existing hypertension | Ischemic stroke (European) | Simple mode | 0.083 | ( 0.029 to 0.137 ) | 0.057 | ✖ | 94 |
| | | Weighted mode | 0.083 | ( 0.031 to 0.134 ) | 0.051 | ✖ | 94 |
| | | Weighted median | 0.079 | ( 0.034 to 0.124 ) | 0.001 | ✔ | 92 |
| | | Inverse variance weighted | 0.060 | ( 0.011 to 0.109 ) | 0.016 | ✔ | 72 |
| Pre-existing hypertension | Cardioembolic IS (Transethnic) | Simple mode | 0.172 | ( 0.064 to 0.280 ) | 0.052 | ✖ | 98 |
| | | Weighted mode | 0.172 | ( 0.067 to 0.277 ) | 0.049 | ✔ | 98 |
| | | Weighted median | 0.172 | ( 0.091 to 0.252 ) | < 0.001 | ✔ | 98 |
| | | Inverse variance weighted | 0.149 | ( 0.084 to 0.214 ) | < 0.001 | ✔ | 94 |
| Pre-existing hypertension | Cardioembolic IS (European) | Simple mode | 0.169 | ( 0.049 to 0.288 ) | 0.070 | ✖ | 95 |
| | | Weighted mode | 0.169 | ( 0.059 to 0.279 ) | 0.057 | ✖ | 95 |
| | | Weighted median | 0.167 | ( 0.079 to 0.254 ) | < 0.001 | ✔ | 94 |
| | | Inverse variance weighted | 0.150 | ( 0.079 to 0.220 ) | < 0.001 | ✔ | 88 |
| Maternal care | Large artery IS (Transethnic) | Simple mode | 0.222 | ( 0.002 to 0.441 ) | 0.186 | ✖ | 75 |
| | | Weighted mode | 0.222 | ( -0.025 to 0.469 ) | 0.220 | ✖ | 75 |
| | | Weighted median | 0.222 | ( 0.016 to 0.427 ) | 0.034 | ✔ | 75 |
| | | Inverse variance weighted | 0.222 | ( 0.052 to 0.392 ) | 0.010 | ✔ | 75 |
| HELLP syndrome | Total stroke (Transethnic) | Simple mode | 0.106 | ( -0.014 to 0.226 ) | 0.097 | ✖ | 100 |
| | | Weighted mode | 0.118 | ( 0.012 to 0.223 ) | 0.040 | ✔ | 100 |
| | | Weighted median | 0.103 | ( 0.049 to 0.158 ) | < 0.001 | ✔ | 100 |
| | | Inverse variance weighted | 0.096 | ( 0.047 to 0.146 ) | < 0.001 | ✔ | 100 |
| HELLP syndrome | Total stroke (European) | Simple mode | 0.079 | ( -0.030 to 0.189 ) | 0.168 | ✖ | 100 |
| | | Weighted mode | 0.088 | ( -0.017 to 0.193 ) | 0.112 | ✖ | 100 |
| | | Weighted median | 0.077 | ( 0.022 to 0.132 ) | 0.006 | ✔ | 100 |
| | | Inverse variance weighted | 0.064 | ( 0.027 to 0.100 ) | 0.001 | ✔ | 100 |
| HELLP syndrome | Ischemic stroke (Transethnic) | Simple mode | 0.061 | ( -0.082 to 0.204 ) | 0.411 | ✖ | 100 |
| | | Weighted mode | 0.083 | ( -0.037 to 0.202 ) | 0.189 | ✖ | 100 |
| | | Weighted median | 0.068 | ( 0.006 to 0.131 ) | 0.032 | ✔ | 100 |
| | | Inverse variance weighted | 0.107 | ( 0.054 to 0.160 ) | < 0.001 | ✔ | 100 |
| HELLP syndrome | Cardioembolic IS (European) | Simple mode | 0.242 | ( -0.020 to 0.503 ) | 0.081 | ✖ | 100 |
| | | Weighted mode | 0.246 | ( 0.023 to 0.468 ) | 0.040 | ✔ | 100 |
| | | Weighted median | 0.172 | ( 0.048 to 0.296 ) | 0.006 | ✔ | 100 |
| | | Inverse variance weighted | 0.137 | ( 0.024 to 0.249 ) | 0.018 | ✔ | 100 |
| Gestational hypertension | Small vessel IS (Transethnic) | Simple mode | 0.181 | ( -0.039 to 0.401 ) | 0.182 | ✖ | 100 |
| | | Weighted mode | 0.183 | ( -0.013 to 0.380 ) | 0.141 | ✖ | 100 |
| | | Weighted median | 0.186 | ( 0.032 to 0.340 ) | 0.018 | ✔ | 100 |
| | | Inverse variance weighted | 0.163 | ( 0.041 to 0.286 ) | 0.009 | ✔ | 100 |
| Gestational hypertension | Small vessel IS (European) | Simple mode | 0.209 | ( -0.053 to 0.471 ) | 0.156 | ✖ | 100 |
| | | Weighted mode | 0.203 | ( -0.047 to 0.453 ) | 0.150 | ✖ | 100 |
| | | Weighted median | 0.197 | ( 0.015 to 0.379 ) | 0.034 | ✔ | 100 |
| | | Inverse variance weighted | 0.211 | ( 0.074 to 0.349 ) | 0.003 | ✔ | 100 |
| Gestational diabetes | Large artery IS (European) | Simple mode | 0.145 | ( -0.003 to 0.293 ) | 0.079 | ✖ | 100 |
| | | Weighted mode | 0.170 | ( 0.058 to 0.282 ) | 0.011 | ✔ | 100 |
| | | Weighted median | 0.197 | ( 0.092 to 0.302 ) | < 0.001 | ✔ | 100 |
| | | Inverse variance weighted | 0.230 | ( 0.125 to 0.334 ) | < 0.001 | ✔ | 100 |
| Excessive vomiting | Ischemic stroke (Transethnic) | Simple mode | 0.082 | ( 0.016 to 0.148 ) | 0.135 | ✖ | 99 |
| | | Weighted mode | 0.082 | ( 0.014 to 0.150 ) | 0.143 | ✖ | 99 |
| | | Weighted median | 0.075 | ( 0.015 to 0.136 ) | 0.014 | ✔ | 97 |
| | | Inverse variance weighted | 0.071 | ( 0.029 to 0.113 ) | 0.001 | ✔ | 95 |
| Excessive vomiting | Ischemic stroke (European) | Simple mode | 0.054 | ( -0.018 to 0.126 ) | 0.281 | ✖ | 56 |
| | | Weighted mode | 0.066 | ( 0.000 to 0.131 ) | 0.189 | ✖ | 73 |
| | | Weighted median | 0.059 | ( 0.000 to 0.118 ) | 0.049 | ✔ | 63 |
| | | Inverse variance weighted | 0.059 | ( 0.010 to 0.108 ) | 0.017 | ✔ | 64 |
